# The use of text mining to obtain a historical overview of research on therapeutic drug monitoring

**DOI:** 10.1101/2023.06.22.23291733

**Authors:** Tetsuo Matsuzaki, Hiroyuki Mizoguchi, Kiyofumi Yamada

## Abstract

Therapeutic drug monitoring (TDM) is a routine clinical practice used to individualize drug dosing and thereby maintain drug efficacy and minimize the consequences of overexposure. TDM is applied to many drug classes, including immunosuppressants, anticoagulants, antineoplastic agents, and antibiotics. Considerable efforts have been made to establish routine TDM practice for each drug. However, because TDM has been developed within the context of specific drugs, there is insufficient understanding of historical trends in TDM research itself.

In this study, we employed text mining approaches to explore trends in the TDM research field. We first performed a PubMed search to determine which drugs and drug classes have been extensively studied in the context of TDM. This investigation revealed that the most commonly studied drugs were warfarin and cyclosporine, followed by tacrolimus, heparin, and vancomycin. In terms of drug classes, most publications focused on antineoplastic agents, anticoagulants, immunosuppressants, and antibiotics. We next used a newly developed python-based module to obtain PubMed records of publications relevant to TDM, and then subjected them to text mining. Our analyses revealed how TDM research has evolved over the years.

## Introduction

Therapeutic drug monitoring (TDM) is a clinical practice used to individualize drug dosing by maintaining serum, plasma, or blood drug concentrations within a therapeutic range. Drug exposure monitoring involves directly measuring serum drug concentrations and monitoring specific drug response markers such as the prothrombin time-international normalized ratio (PT-INR) for warfarin [1–4]. The past 30 years have seen tremendous advances in TDM research, which has resulted in increased adoption of TDM for a broad range of drugs [2,5–7]. Due to the nature of research on drugs, TDM research has been conducted in the context of specific drugs; however, this poses a challenge to our comprehensive understanding of the trends in the TDM research field. Indeed, there have been few investigations and reviews of this field.

There has been a dramatic increase in the number of publications in all fields, not only that of TDM research. Because of the vast amount of published literature, researchers can no longer read all available articles, even in their own narrow discipline. This challenge can be tackled by text mining techniques [8]. Text mining is the process of extracting and processing text to derive meaningful insight from textual data. It makes it possible to perform targeted navigation of the knowledge landscape, thus helping researchers to guide their endeavors. Studies applying text mining techniques have been increasingly reported [9–11]. Recently, web-based platforms to facilitate text mining analysis have been developed. For example, Wei et al. developed PubTator, a web-based system that assists in literature curation [12,13]. More recently, Chen et al. developed LitCovid, a curated literature hub dedicated to coronavirus disease 2019 (COVID-19) that enables tracking the latest COVID-19 research [14].

In the present study, we sought to derive a comprehensive overview of trends in TDM research by analyzing publications indexed in PubMed (https://pubmed.ncbi.nlm.nih.gov). We first determined which drugs and drug classes have been extensively studied in the context of TDM by analyzing the numbers of relevant publications. We also developed a python module that enables the automated extraction of titles and abstracts of selected articles by PubMed ID. Using this module, we obtained a dataset comprising records of publications relevant to TDM and subjected it to text mining. These analyses uncovered longitudinal trends in TDM research.

## Materials and Methods

### Python modules

The python modules used in this study are available at https://github.com/Matsuzaki-T/TDM_textmining. Retrieval of drug names from KEGG DRUG Drugs deposited in the KEGG DRUG Database (https://www.genome.jp/kegg/drug/) were analyzed in this study [15]. Extraction of records from KEGG DRUG was performed on March 30, 2023. Entry IDs, drug names, and drug efficacies were automatically extracted using python module 1.0.

### Measuring the number of TDM-relevant publications on each drug

The drug list collected by module 1.0 was subjected to module 2.0, which resulted in a list that included the Pubmed IDs of TDM-related publications for each drug. Of the entries in KEGG DRUG, we excluded human serum and compounds that are not considered to be drugs (e.g., water), or that are not administered for treatment (e.g., alcohol) (Supplemental Table 1).

To investigate which drug classes (determined by efficacies in KEGG DRUG) were well-studied in the context of TDM, the drug list obtained by module 2.0 was subjected to module 2.1.

Based on the number of TDM-relevant publications, we grouped drugs into 3 groups: the major group (100 or more publications), the moderate group (10–99 publications), and the minor group (fewer than 10 publications). We investigated the publications in each group and used module 2.2 to visualize the results in the form of a Venn diagram.

### Topic word extraction by word cloud

To perform automated text mining, we first attempted to retrieve text data from PubMed. easyPubMed (https://cran.r-project.org/web/packages/easyPubMed/index.html) is a readily available R-interface that enables automated extraction of PubMed records. However, it was recently shown that easyPubMed yields fewer results than the PubMed website (reported in https://github.com/dami82/easyPubMed/issues/4). Thus, we developed a new interface that accurately parses PubMed documents and retrieves their contents. Module 3.0 enables the retrieval of titles and abstracts of selected articles from PubMed ID by accessing PubTator Central (PTC), which stores and annotates the abstracts of all PubMed articles [12,13]. The titles and abstracts of articles stored in PTC are easily retrieved by queries using the characters “|t|” and “|a|” in search results, respectively (a detailed description is presented on the PTC website, https://www.ncbi.nlm.nih.gov/research/pubtator/api.html). A list of PubMed IDs was obtained via the PubMed website using the search term “(therapeutic drug monitoring [MeSH Terms]) AND (drug name [MeSH Terms]) AND (“1900/1/1”[Date - Publication]: “2022/12/31”[Date - Publication]),” and was subjected to module 3.0 on March 30, 2023, resulting in a list of the titles and abstracts of 23,393 TDM-related publications. However, when investigating the output of the module, we noticed that 5 of 23,393 publications were not stored in PTC, and therefore the records of these publications were added manually.

Records were divided into 4 groups based on publication date: period I (before 2004), period II (2004–2011), period III (2012–2016), and period IV (2017–2022). These periods were defined so that the number of publications was similar in each (approximately 6,000 publications per period). We retrieved biomedical entities from each title using the spaCy model “en_core_sci_lg,” which has been trained on biomedical text and enables the recognition of biomedical entities in documents [16]. We removed stop words, defined as entities that are commonly used in TDM research (e.g., therapy, monitoring, drug, and effect). The complete list of stop words is available in Supplemental Table 3. The obtained list of biomedical entities was then subjected to word cloud (module 4.0), yielding a visual representation of the topic words.

### Latent Dirichlet Allocation (LDA)

LDA requires a document-term matrix as an input data structure [17]. Therefore, we began converting the abstracts of the publications obtained by module 3.0 into a document-term matrix. We retrieved biomedical entities from each abstract as in the word cloud analysis. Then, the obtained document-term matrix was converted into a Bag-of-Words representation by CountVectorizer from sklearn (https://scikit-learn.org/stable/modules/generated/sklearn.feature_extraction.text.CountVectorizer.html). The resultant Bag-of-Words matrix was subjected to LDA using the LDA module from sklearn (https://scikit-learn.org/stable/modules/generated/sklearn.decomposition.LatentDirichletAllocation.html). We conducted LDA with different numbers of topics, ranging from 10 to 20. These analyses were conducted using module 5.0.

## Results

### The number of publications in TDM research fields

In this study, we analyzed PubMed articles with publication dates before 2023. We first investigated all publications relevant to TDM and examined changes in the number of publications over time. A PubMed search using the term “(therapeutic drug monitoring [MeSH Terms]) AND (“1900/1/1”[Date - Publication] : “2022/12/31”[Date - Publication])” yielded 23,393 publications. The earliest publications, which reported the method used to estimate serum concentrations of streptomycin and penicillin, appeared in 1948 [18]. Figure 1 shows the number of TDM-related publication between 1940 and 2022. The number of publications gradually increased until peaking at 1,417 publications in 2016, then decreased to 445 publications in 2022, a number almost equivalent to that in the mid-1990s.

**Figure 1.**
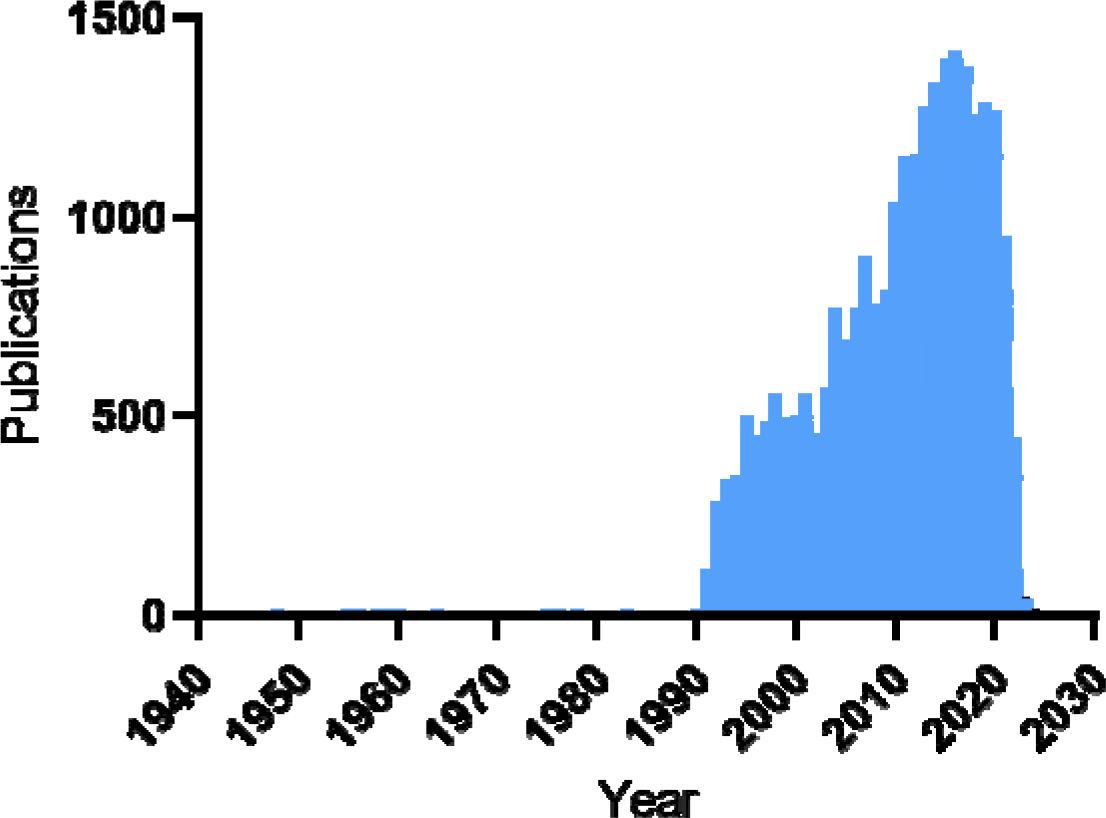
The number of TDM-related publications between 1948 and 2022.

### Investigating the number of TDM-related publications for each drug

We next investigated which drugs and drug classes have been extensively studied in the context of TDM. Toward this end, we retrieved drug information from the KEGG DRUG Database (https://www.genome.jp/kegg/drug/), a comprehensive information resource for drugs approved in Japan, the United States (U.S.), and Europe [15]. Extraction of the KEGG DRUG records (i.e., drug ID, drug names, and drug classes) were conducted using python-based module 1.0 (Figure 2A). A PubMed search was performed for each selected drug name using the following search term: “(drug name [MeSH Terms]) AND (therapeutic drug monitoring[MeSH Terms]) AND (“1900/1/1”[Date - Publication] : “2022/12/31”[Date - Publication]).” The PubMed search was conducted in an automated manner using python-based module 2.0 (Figure 2A).

**Figure 2.**
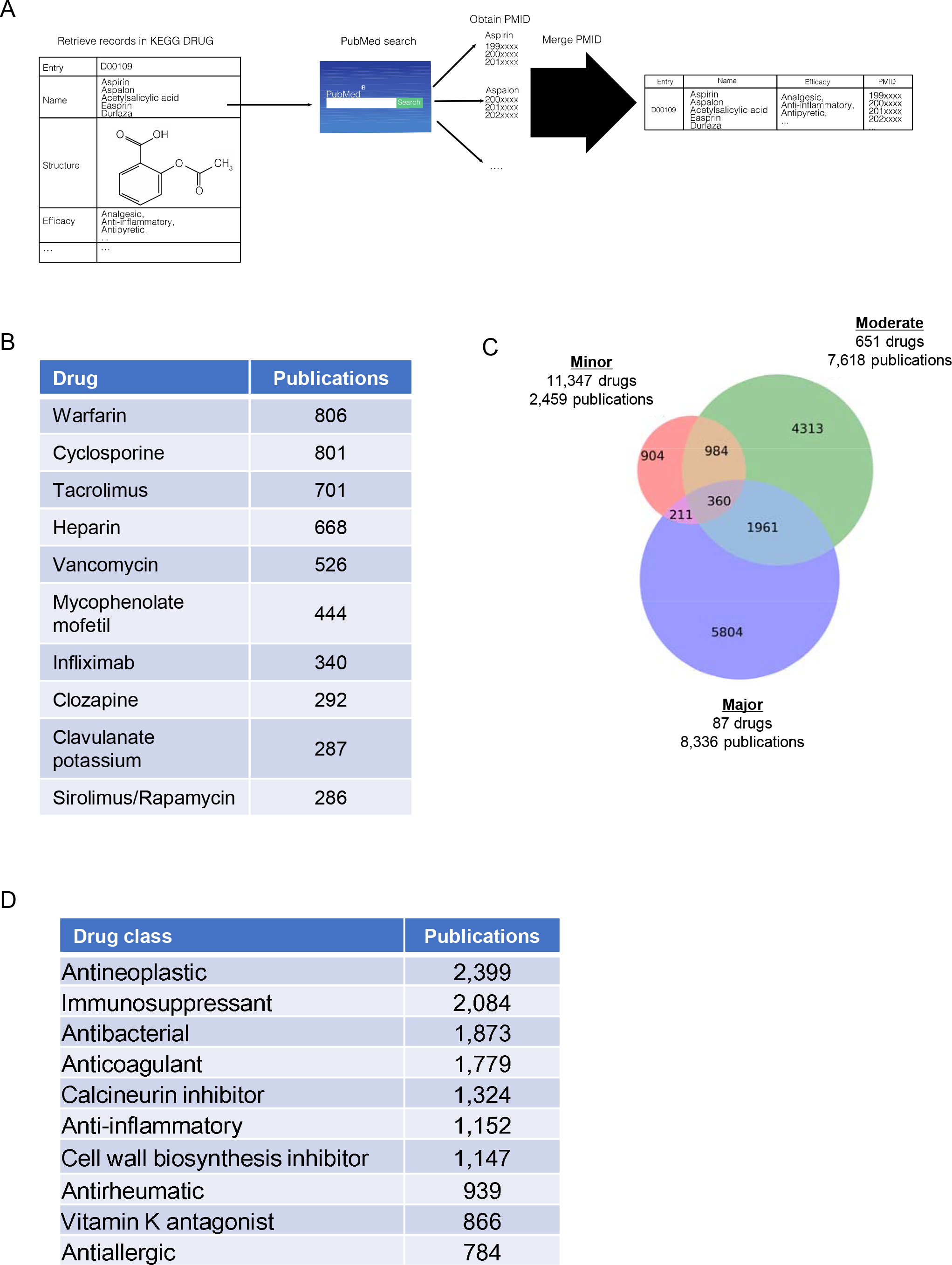
(A) A schematic of the extraction of TDM-related publications on each drug. Each drug name in the name column was subjected to a PubMed search, and all resulting PubMed IDs (PMIDs) were then merged. (B) The top 10 most studied drugs in the context of TDM. (C) Venn diagrams of TDM-related publications in each drug group. (D) A ranking of drug classes in terms of the number of TDM-related publications.

A PubMed search showed that “globulin, immune” (D06458) was the top-ranked drug in terms of number of publications (Supplemental Table 1). However, a PubMed search with the term “(globulin, immune [MeSH Terms]) AND (therapeutic drug monitoring [MeSH Terms]) AND (“1900/1/1”[Date - Publication] : “2022/12/31”[Date - Publication])” yielded articles relevant to antibody therapies encompassing a broad range of diseases (e.g., infliximab, rituximab, and nivolumab, data not shown). Thus, we excluded “globulin, immune” from this ranking and regarded warfarin (806 publications) and cyclosporine (801 publications) as the most frequently studied drugs regarding TDM, followed by tacrolimus (701 publications), heparin (668 publications), and vancomycin (526 publications) (Figure 2B and Supplemental Table 1).

We classified drugs into 3 groups based on number of publications: the major group (100 or more publications), the moderate group (10–99 publications), and the minor group (fewer than 10 publications). Based on drug ID, the numbers of drugs categorized into the major, moderate, and minor groups were 87, 651, and 11,347, respectively (Supplemental Table 1). It should be noted that there was overlap of drugs in KEGG DRUG; for example, D00564, D01280, and D08682 all indicate warfarin. However, because applying manual data cleaning to such a large dataset was not practical, we counted the publications on each drug based on drug ID. There were 8,336 publications on drugs in the major group, accounting for 35.6% of the total of 23,393 publications. There were 7,618 (32.6%) and 2,459 (10.5%) publications on drugs in the moderate and minor groups, respectively (Figure 2C). This finding indicates that a small number of extensively studied drugs accounted for a large proportion of TDM-related publications. Of note, only 62.1% (14,537/23,393) of TDM-related publications were recovered by drug name search (Figure 2C). This indicates that a large number of TDM-related publications have not been assigned a drug name MeSH term.

When we focused on drug classes (corresponding to efficacies in KEGG DRUG), the most studied class was antineoplastic (hereinafter, antineoplastic agents), followed by immunosuppressants, antibacterials (hereinafter, antibiotics), anticoagulants, and calcineurin inhibitors (Figure 2D). While immunosuppressants, anticoagulants, and antibiotics contained drugs that ranked in the top 5 in terms of number of publications, antineoplastic agents did not: sirolimus/rapamycin was the most studied of these drugs, and ranked 10th in the number of publications (Figure 2B and Supplemental Table 1). This indicates that TDM studies have been conducted on a broad range of antineoplastic agents, thereby resulting in the largest number of publications for this drug class. Indeed, antineoplastic agents were the predominant drug type in the major group; this group contained 11 antineoplastic agents, and 5, 7, and 5 anticoagulants, immunosuppressants, and antibiotics, respectively (Supplemental Table 2).

### Visualization of historical changes in topic words in TDM research

We next analyzed how themes of TDM research have changed over time. To this end, we first extracted text data (i.e., titles and abstracts) from PubMed, then subjected them to text mining analysis. easyPubMed is an R-based interface that enables automated extraction of PubMed records (https://cran.r-project.org/web/packages/easyPubMed/index.html). However, it has been reported that fewer PubMed records are obtained via easyPubMed than from the PubMed website (reported in https://github.com/dami82/easyPubMed/issues/4). To overcome this problem, we developed a python-based program to accurately retrieve titles and abstracts of selected publications (Figure 3A). Using this program, we retrieved the titles and abstracts of 23,393 publications. We classified publications into 4 groups based on publication date: period I (before 2004), period II (2004–2011), period III (2012–2016), and period IV (2017–2022). These were defined so that the number of publications was evenly distributed (approximately 6,000 publications per period). These periods also corresponded to the emergence of the TDM research field (period I), the initial rise in the number of publications (period II), a further rise in publication number with a peak in 2016 (period III), and a decline in publication number (period IV).

**Figure 3.**
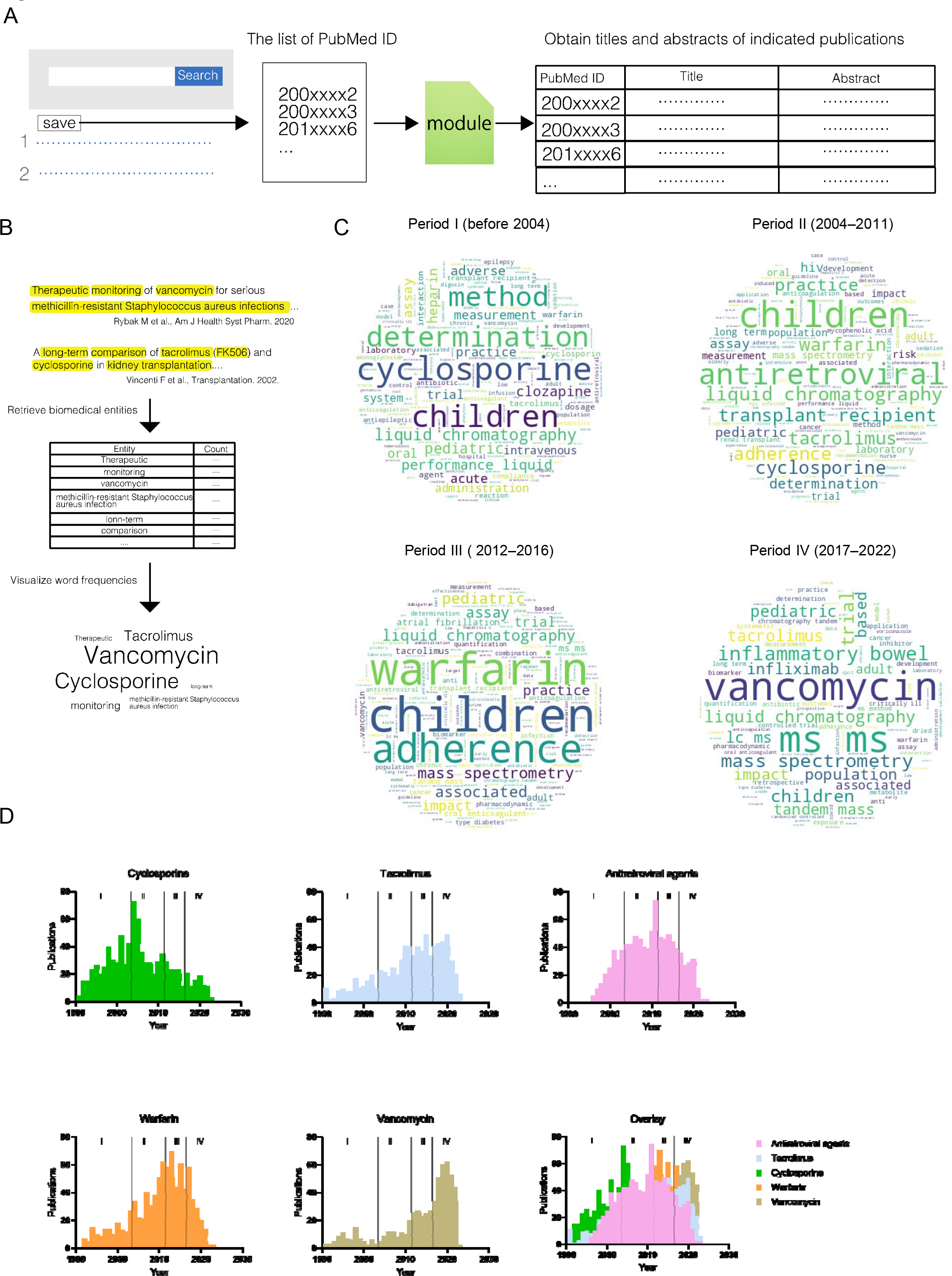
(A) An overview of the pipeline for retrieving the titles and abstracts of indicated publications by PubMed ID. (B) A schematic of word cloud analysis. (C) Word cloud representation of topic words in each period. (D) The number of TDM-related publications for the indicated drugs between 1990 and 2022. The data of each drug are displayed in a single histogram (“Overlay”). I, II, III, and IV indicate periods I, II, III, and IV, respectively.

We next retrieved topic words in each period and visualized them using word cloud, a visualization module used to depict the frequencies of words in documents (Figure 3B) [19]. We extracted biomedical entities from the title of each publication by leveraging the spaCy model “en_core_sci_lg,” which was trained using biomedical text [16]. Figure 3C shows the word cloud representation of the topic words in each period. “Children” and terms relevant to analytical methods, such as “liquid chromatography” (LC) and “mass spectrometry” (MS), were major topic words across all periods. The prevalence of the term “children” indicates that pharmacokinetics and pharmacodynamic in children have been extensively studied. The latter 2 terms reflected studies that applied established LC or MS methods or developed new ones, in both cases to monitor drug concentrations [6,20–22]. Word cloud also identified topic words that were distinct to each period, as follows. In period I, “cyclosporine” was the most common topic word, followed by “tacrolimus.” In period II, “tacrolimus” was as common as “cyclosporine.” We speculated that accumulating evidence of tacrolimus as a valuable therapeutic alternative to cyclosporine from the 1990s to early 2000s stimulated the increase in associated TDM studies [23–27]. “Antiretroviral” emerged as a major topic word in period II. To determine the reason for this, we conducted a word cloud analysis of publications obtained using the following search term: “(antiretroviral agents [MeSH Terms]) AND (therapeutic drug monitoring [MeSH Terms]) AND (“2004/1/1”[Date - Publication] : “2011/12/31”[Date - Publication]).” This search retrieved a broad range of drugs targeting human immunodeficiency virus (HIV) that were developed after the late 1990s (Supplemental Figure 1) [21,28]. Thus, we speculated that the predominance of the term “antiretroviral” reflected remarkable progress in anti-HIV agents. “Warfarin” was another major topic word in period II, and its frequency further increased in period III, when it became one of the most predominant topic words. This is presumably due to the emergence of direct oral anticoagulants (DOACs) such as dabigatran and rivaroxaban after the late 2000s, which stimulated comparative studies of DOACs and warfarin [29–31]. Period IV was marked by the predominance of the topic word “vancomycin.” A plausible explanation for this finding is that the first consensus guideline for vancomycin TDM was published in 2009, and this facilitated studies evaluating this guideline [32–35]. Indeed, due to the accumulation of evidence after 2009, a revised guideline was published in 2020 [2]. Terms associated with inflammatory bowel disease (IBD), such as “inflammatory bowel” and “infliximab,” were also featured in period IV, indicating that TDM of drugs for IBD was extensively studied during this time [36,37].

Topic words retrieved by word cloud analysis in each period largely coincided with the number of publications: TDM-related publications on antiretroviral agents, warfarin, and vancomycin peaked in periods II, III, and IV, respectively (Figure 3D).

### Topic model analysis by LDA

Lastly, we attempted to categorize publications based on their abstracts. Toward this end, we applied LDA, which is well-established technique for performing topic modeling [17]. With LDA, each document can be tagged with topics, making it possible to classify documents based on their content. Figure 4A shows a schematic diagram of the analysis. We started by converting each abstract into a matrix of token counts. We extracted biomedical entities from the abstract of each publication as in Figure 3. The obtained collection of biomedical entities was converted into a Bag-of-Words representation [17]. Bag-of-Words is a method of representing text that describes the frequency of words within a document. For each publication, we obtained an abstract-entities matrix by counting the number of occurrences of each entity in the abstract, and then subjected the matrix to LDA. First, we determined the optimal number of topics for LDA, between 10 and 20, by measuring perplexity. Perplexity is an indicator of the predictive power and generalizability of a topic model, with a lower perplexity indicating better generalization ability [17]. Perplexity decreased as the number of topics increased, and reached a minimum when the number of topics was 19 (Supplemental Figure 2A). Based on this result, we set the number of topics for LDA analysis to be 19.

**Figure 4.**
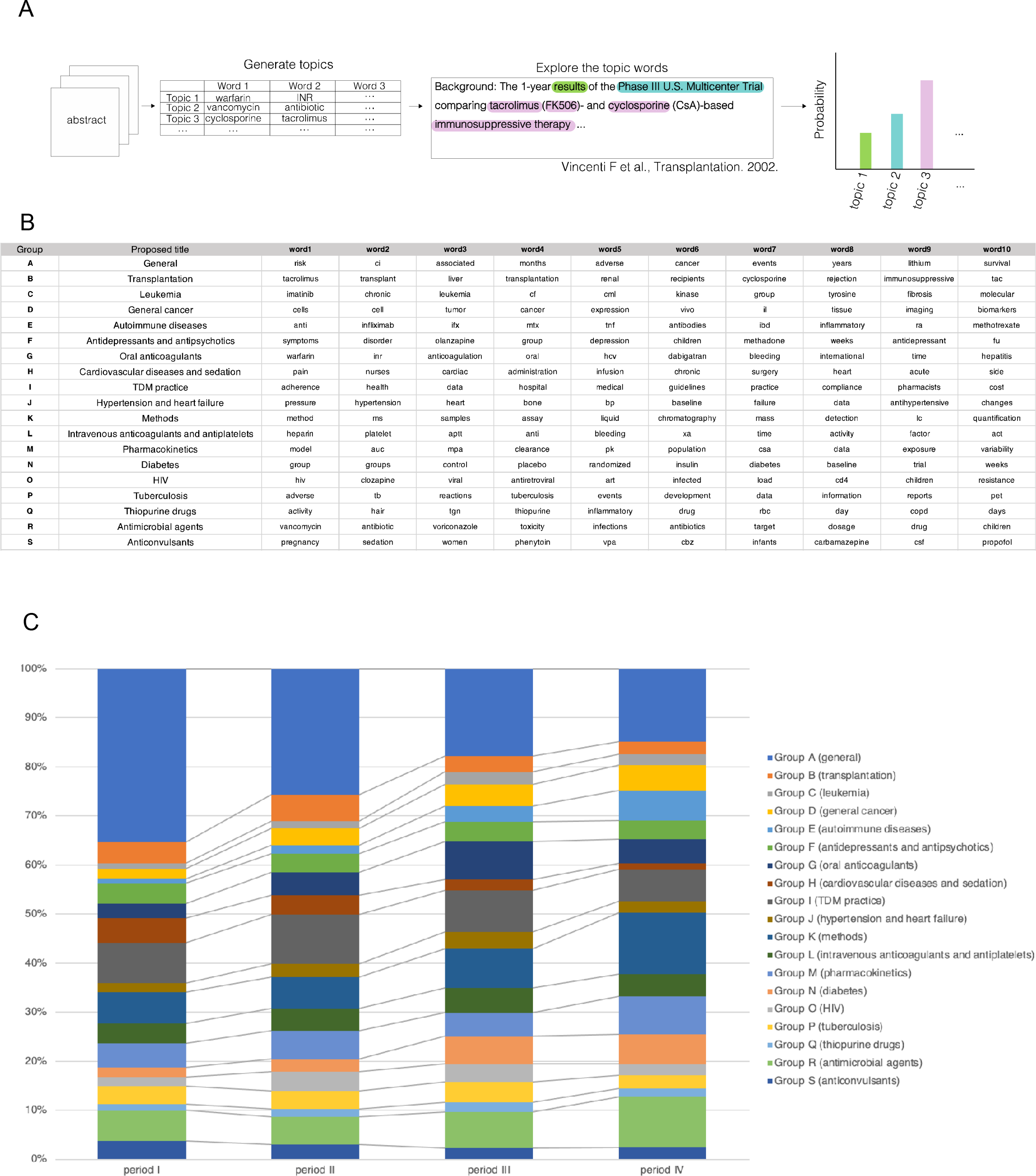
(A) A schematic of LDA. (B) Proposed titles and the top 10 terms of each topic. (C) Change in the percentage of publication counts of each topic.

We listed group names alphabetically, with the title of each group derived based on the topic words retrieved by LDA and word cloud analysis (Figure 4B, Supplemental Figure 2B, 3, and Supplemental Table 4). While we assigned most groups a distinct title, we could not do so for group A due to the presence of multiple topic words, specifically warfarin, cyclosporine, HIV, and vancomycin, with unclear relationships between them (Supplemental Figure 2B). We speculated that the publications not categorized into other groups were included in group A. Likewise, group H (cardiovascular disease and sedation) contained topic words with ambiguous connections. We speculated that publications relevant to cardiovascular disease and sedation were grouped together because they were associated with the treatment of critically ill patients [38,39]. Of note, while group D (general cancer) included publications relevant to cancer, those regarding leukemia constituted a discrete group (group C). The word cloud representation of group C featured terms relevant to kinase inhibitors (e.g., “kinase inhibitor,” “tyrosine kinase,” and “imatinib”) and the indications for these drugs (e.g., “chronic myeloid” and “myeloid leukemia”) (Supplemental Figure 2B). These results indicate that TDM research in the cancer field has focused heavily on the use of kinase inhibitors for the treatment of chronic myeloid leukemia.

Across all periods, group A (general) contained the largest number of publications (Figure 4C), although this number trended downward after period I. Likewise, group I (TDM practice), group K (methods), and group R (antibiotics and antifungals) contained large numbers of publications across all periods. Regarding group I, this indicates that sustained efforts have been made to establish the clinical significance of TDM practice. In terms of group K, the results in all periods were consistent with the word cloud analysis, which featured methodological terms (e.g., liquid chromatography and mass spectrometry) (Figure 3C). Concerning group R, word cloud featured vancomycin across all periods, indicating that vancomycin was the central theme of TDM pertaining to antibiotics and antifungals (Supplemental Figure 3R). Voriconazole was also featured in group R beginning in period II, which largely coincides with the U.S. approval of voriconazole in 2002.

In period I, group A (general), group I (TDM practice), group K (methods), and group R (antibiotics and antifungals) contained the largest numbers of publications, specifically 2,003 (35.4%), 468 (8.3%), 363 (6.4%), and 355 (6.3%), respectively (Figure 4C and Supplemental Table 4). In period II, group B (transplantation), group G (oral anticoagulants), group I (TDM practice), and group O (HIV) showed increased numbers of publications relative to period I: from 242 (4.3%) to 350 (5.4%), 169 (3.0%) to 305 (4.7%), 468 (8.3%) to 652 (10.1%), and 109 (1.9%) to 251 (3.9%), respectively (Figure 4C and Supplemental Table 4). For groups B, G, and O, these findings are consistent with the results of the word cloud analysis, which showed that “cyclosporine,” “tacrolimus,” “warfarin,” and “antiretroviral” were common words in period II (Figure 3C). For group I, we further subjected the publications from period II to word cloud analysis. “Antiretroviral” and “adherence” were 2 of the most frequent words in the word cloud visualization (Supplemental Figure 3I). This indicated that studies on HIV treatment adherence were conducted during period II, and therefore the increased number of group I publications relative to period I was partly associated with the increased number of publications in group O (HIV) [40,41].

In period III, group G (oral anticoagulants) showed further increases in the number of publications compared to period II, from 305 (4.7%) to 441 (7.7%) (Figure 4C and Supplemental Table 4). This was consistent with the results of the word cloud analysis in period III, in which “warfarin” was the most common topic word (Figure 3C). In this period, the number of publications in group N (diabetes) also increased relative to period II, from 167 (2.6%) to 322 (5.6%) (Figure 4C and Supplemental Table 4). We subjected the group N publications in period III to word cloud analysis and found that words relevant to clinical trials were featured (e.g., “controlled trial,” “randomized,” “blind,” and “placebo”; Supplemental Figure 3N). This indicates that a large number of clinical trials on diabetes were conducted in this period [42,43]. Publications related to cancer (i.e., groups C and D) increased in number between period I and period III (Figure 4C and Supplemental Table 4). While the number of publications in group C (leukemia) peaked in period III, the number in group D continued to increase. These trends were consistent with the numbers of publications on TDM research in the cancer field (Supplemental Figure 4).

In period IV, group E (autoimmune diseases), group K (methods), group M (pharmacokinetics), and group R (antibiotics and antifungals) demonstrated increased numbers of publications compared to period III: from 181 (3.2%) to 341 (6.2%), 455 (8.0%) to 696 (12.6%), 278 (4.9%) to 431 (7.8%), and 425 (7.4%) to 570 (10.3%), respectively (Figure 4C and Supplemental Table 4). The increase seen in group E (autoimmune diseases) was largely attributed to publications on IBD, because word cloud analysis featured associated topic words (e.g., “inflammatory bowel,” “infliximab,” “adalimumab,” and “vedolizumab”) (Supplemental Table 3E). The increase in the number of publications on IBD in period IV was consistent with the results of word cloud analysis shown in Figure 3C. For group K (methods), word cloud analysis identified nonspecific words related to analytical methods, making it difficult to determine which topics were most relevant (Supplemental Figure 3B). Thus, we removed these words as stop words and again conducted word cloud analysis (the stop words in this new analysis are available in Supplemental Table 5). Once again the analysis failed to identify characteristic topic words in this period (Supplemental Figure 4T). These results indicate that in period IV, methods-related TDM research was conducted on a broad range of drugs. In period IV, the increased number of publications in group R (antibiotics and antifungals) coincided with the increase in publications on vancomycin TDM (Figure 3D). For group M (pharmacokinetics), we conducted word cloud analysis and found that vancomycin was one of the most featured words (Supplemental Table 3M). Therefore, the increased number of publications in group M was associated with the increase in group R (antibiotics and antifungals). We propose a historical overview of TDM research in Figure 5.

**Figure 5.**
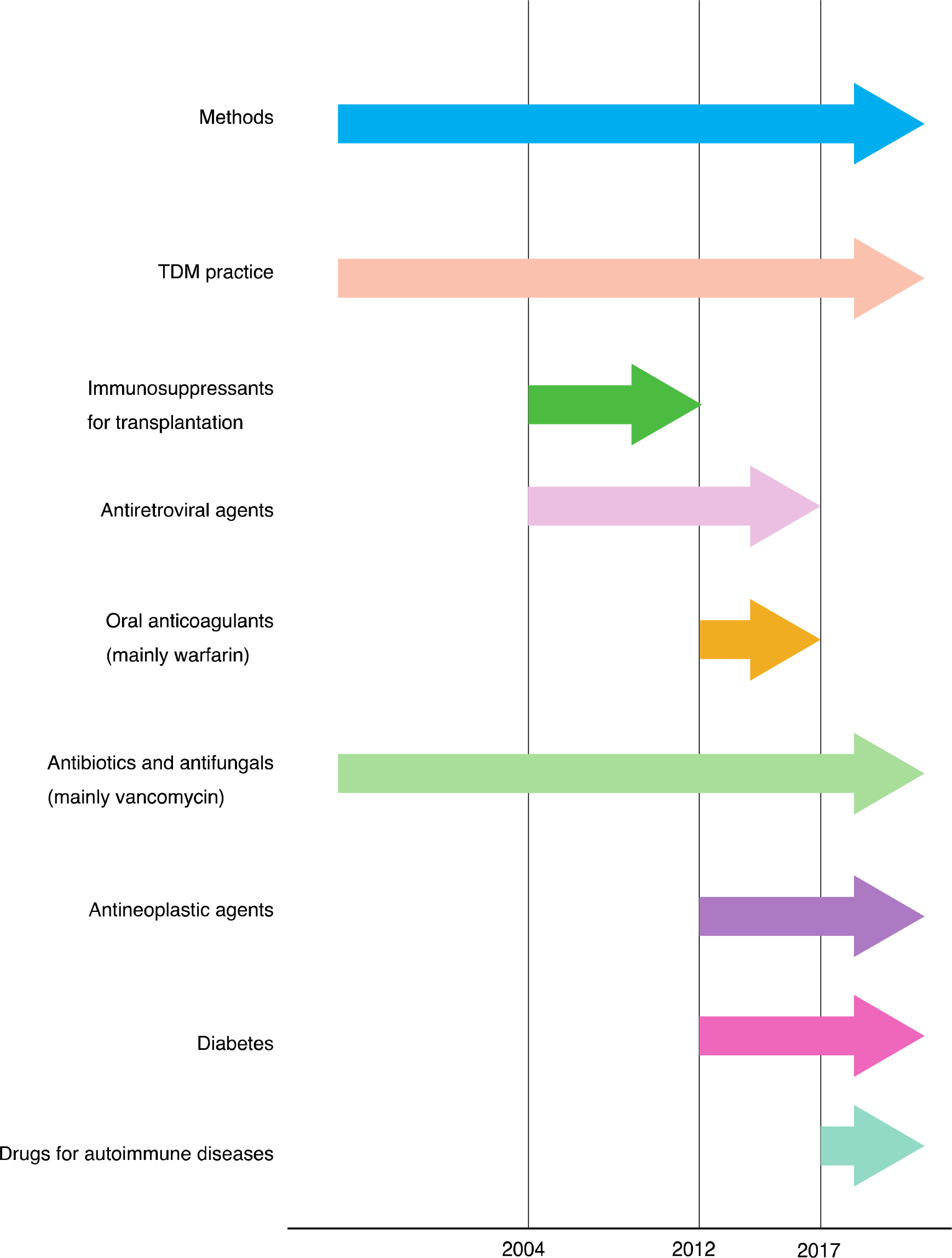
Evolution of the TDM research field. Major themes in each period are described.

## Discussion

Text mining techniques have been employed in many research fields to retrieve useful information from unstructured textual data [9–11]. To our knowledge, this is the first study that employs text mining techniques to provide a historical overview of TDM research trends.

The number of publications relevant to TDM increased until 2016, and decreased thereafter (Figure 1). This indicates a downward trend in TDM research activity. By applying an automated system to perform PubMed searches, we found that according to the number of publications, warfarin and cyclosporine were the most studied drugs in the context of TDM, followed by tacrolimus and heparin (Figure 2B). Among antimicrobial agents, vancomycin was the top-ranked drug, as expected based on its clinical importance and widespread use in clinical settings; a previous analysis of 576 U.S. hospitals reported that vancomycin was one of the most used antibiotics in 2016 [44]. When drugs were classified according to the numbers of related publications, those with publication counts ≥ 100 (the major group) represented fewer than 1% of drugs in KEGG DRUG (Figure 2C and Supplemental Table 1). This indicates that a very small number of drugs have been the focus of most TDM research. It is interesting that the most studied drug class was antineoplastic agents, even though top-ranked drugs such as warfarin, cyclosporine, and vancomycin were not included in this class (Figure 2B and 2D). This indicates that a wide variety of antineoplastic agents were studied in the context of TDM (Supplemental Table 2).

We next adopted well-established text mining techniques to determine how the TDM research field evolved over time. To this end, we first developed a novel python-based module to retrieve titles and abstracts of indicated articles, because the existing interface for querying PubMed records leads to inaccurate results. While testing this module, we noticed that a small number of TDM-related publications that were indexed in PubMed were not indexed in PTC (5 of 23,393 publications; data not shown). Because the module detects such publications and returns the PubMed IDs, users can manually add records to the module output.

The result of word cloud analysis identified distinct topic words in each period: “cyclosporine” and “tacrolimus” in period I; “antiretroviral,” “cyclosporine,” and “tacrolimus” in period II; “warfarin” in period III; and “vancomycin” in period IV (Figure 3C). “Children” and terms relevant to methods (e.g., “liquid chromatography” and “mass spectrometry”) were featured in all periods, indicating that sustained efforts have been made to establish and improve TDM of drugs used in pediatric populations and the methods for monitoring drug concentrations. The topic words retrieved by word cloud analysis were largely consistent with those from LDA (Figure 4B and 4C).

A historical overview of TDM research is summarized in Figure 5. In period I (before 2004), TDM research centered on methods of measuring drug concentrations, TDM practice, and TDM of antibiotics. These topics remained important across all periods. The main topics of publications in period II (2004–2011) were immunosuppressants and anti-HIV agents. TDM of antineoplastic agents was one of the major topics in period III (2012–2016) (Supplemental Table 4). Period III also saw increases in TDM-related publications on anticoagulants (mainly warfarin). The surge in publications on warfarin reflects the emergence of DOACs, which stimulated research comparing DOACs to warfarin [29–31]. Finally, period III was characterized by an increase in publications on TDM of diabetes-related drugs, which was associated with the large number of clinical trials in diabetes during this time [42,43]. In period IV (2017–2022), there was a high proportion of studies on TDM of antibiotics and antifungals, mainly due increased research on vancomycin (Figure 3C). TDM studies related to IBD showed a steady increase across periods and became one of the main topics in period IV (Figure 4C and Supplemental Table 4).

The limitations of this study were as follows. First, we retrieved prescription drugs from the KEGG DRUG Database, but as with other drug databases, KEGG DRUG is not comprehensive. While we annotated drug classes based on the records in KEGG DRUG, these records were sometimes inconsistent with those used in clinical settings. For instance, during the preparation of this manuscript, thalidomide (D00754) was recorded as antibiotic and antineoplastic agent, even though it is not used as an antibiotic in clinical settings [45,46]. Second, we analyzed only publications recorded in PubMed. Evaluating publications recorded in other repositories such as Google Scholar (https://scholar.google.com) and Web of Science (https://clarivate.com/webofsciencegroup/solutions/web-of-science/) will provide a more accurate and comprehensive historical overview of TDM research. Third, we queried TDM-related publications based on MeSH indexing, and therefore this study excluded publications not assigned MeSH terms. Indeed, only 62.1% (14,537/23,393) of TDM-related publications were recovered by drug name search, indicating that large numbers of publications were not assigned MeSH terms for drug names (Figure 2C).

Collectively, we used text mining tools to provide an overview of how the TDM research field has evolved over time, thereby serving as a foundation for future studies.

## Data Availability

All data produced are available online at https://github.com/Matsuzaki-T/TDM_textmining

## Acknowledgements

This work was supported by JSPS KAKENHI (Grant Numbers JP 20H03428, 22K19749, and 23H02669).

## Supplementary material

Supplemental material is available online only.

Code for the modules used to derive the results in this study is available at https://github.com/Matsuzaki-T/TDM_textmining.

---

**Supplemental Figure 1**

Word cloud representation of topic terms in publications relevant to antiretroviral agents in period II.

**Supplemental Figure 2**

(A) The perplexity of LDA models with different numbers of topics. (B) Word cloud representation of topic terms in each group.

**Supplemental Figure 3**

(A–S) Word cloud visualization was conducted for the indicated groups and periods.

(T) A modified list of stop words (Supplemental Table 5) was used for entity extraction, then topic words were visualized as in A–S.

**Supplemental Figure 4**

The number of TDM-related publications for antineoplastic agents. The publication records were retrieved using the following search term: “(therapeutic drug monitoring [MeSH Terms]) AND (antineoplastic agents [MeSH Terms]) AND (“1900/1/1”[Date - Publication]: “2022/12/31”[Date - Publication])”.

