## Supplemental Figures for "The use of text mining to obtain a historical overview of research on therapeutic drug monitoring"

### Slide 1
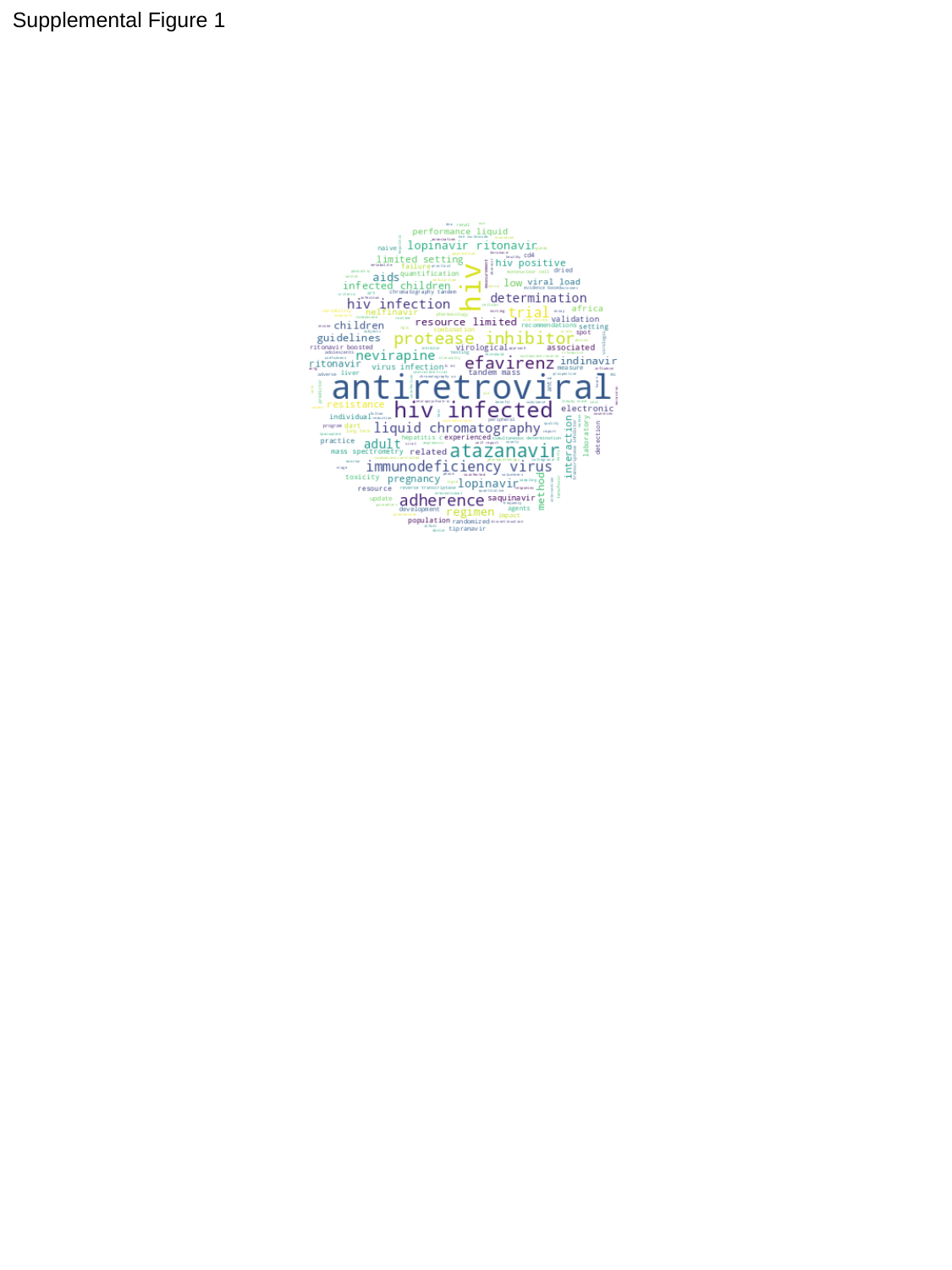

Supplemental Figure 1

### Slide 2
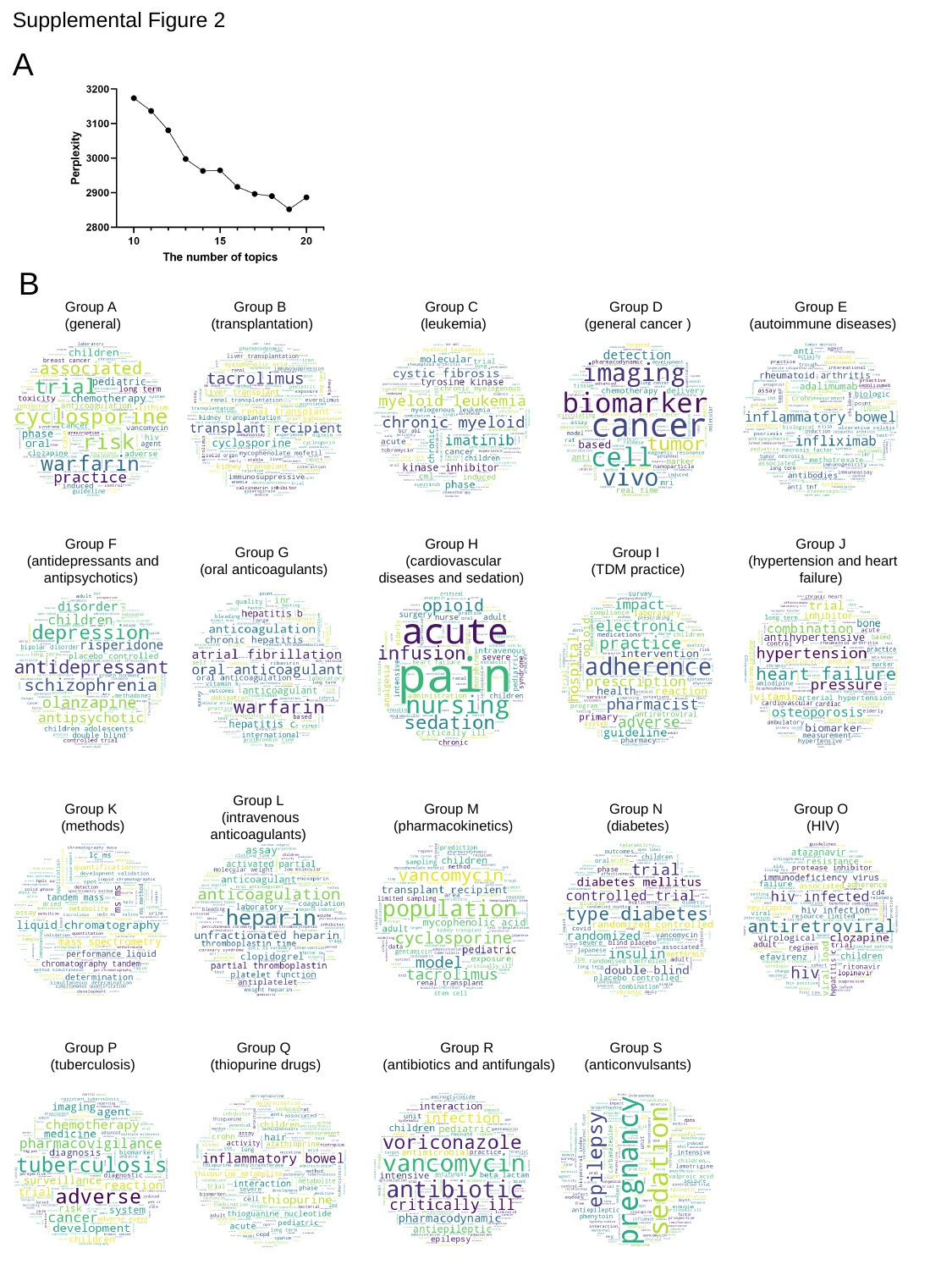

Supplemental Figure 2
A
B
Group A
 (general)
Group B
 (transplantation)
Group C
 (leukemia)
Group D
 (general cancer )
Group E
 (autoimmune diseases)
Group F
 (antidepressants and antipsychotics)
Group H
 (cardiovascular diseases and sedation)
Group J
 (hypertension and heart failure)
Group G
 (oral anticoagulants)
Group I
 (TDM practice)
Group L
 (intravenous anticoagulants)
Group K
 (methods)
Group M
 (pharmacokinetics)
Group N
 (diabetes)
Group O
 (HIV)
Group R
 (antibiotics and antifungals)
Group P
 (tuberculosis)
Group Q
 (thiopurine drugs)
Group S
 (anticonvulsants)

### Slide 3
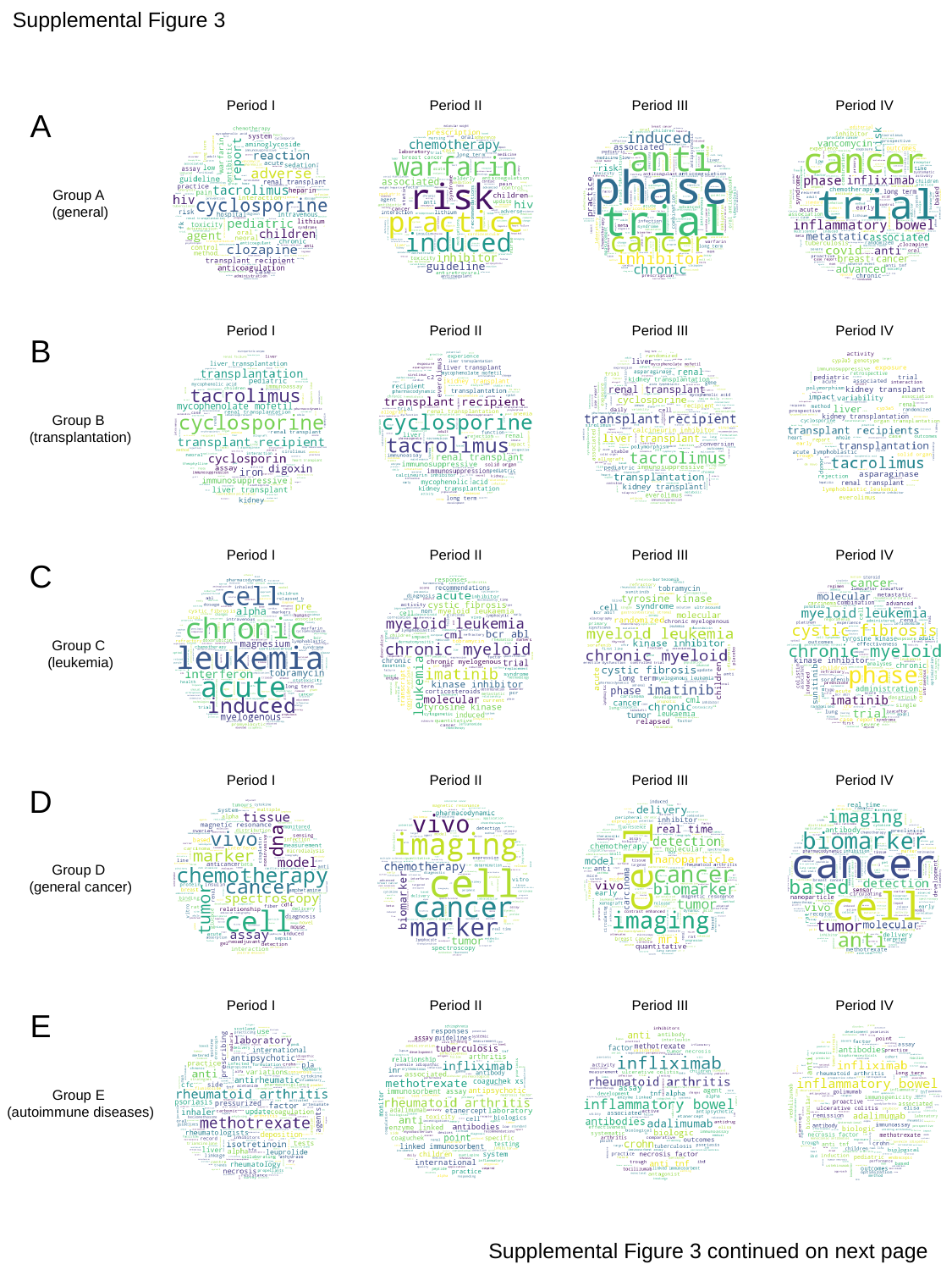

Supplemental Figure 3
Period I
Period II
Period III
Period IV
A
Group A
 (general)
Period I
Period II
Period III
Period IV
B
Group B
 (transplantation)
Period I
Period II
Period III
Period IV
C
Group C
 (leukemia)
Period I
Period II
Period III
Period IV
D
Group D
 (general cancer)
Period I
Period II
Period III
Period IV
E
Group E
 (autoimmune diseases)
Supplemental Figure 3 continued on next page

### Slide 4
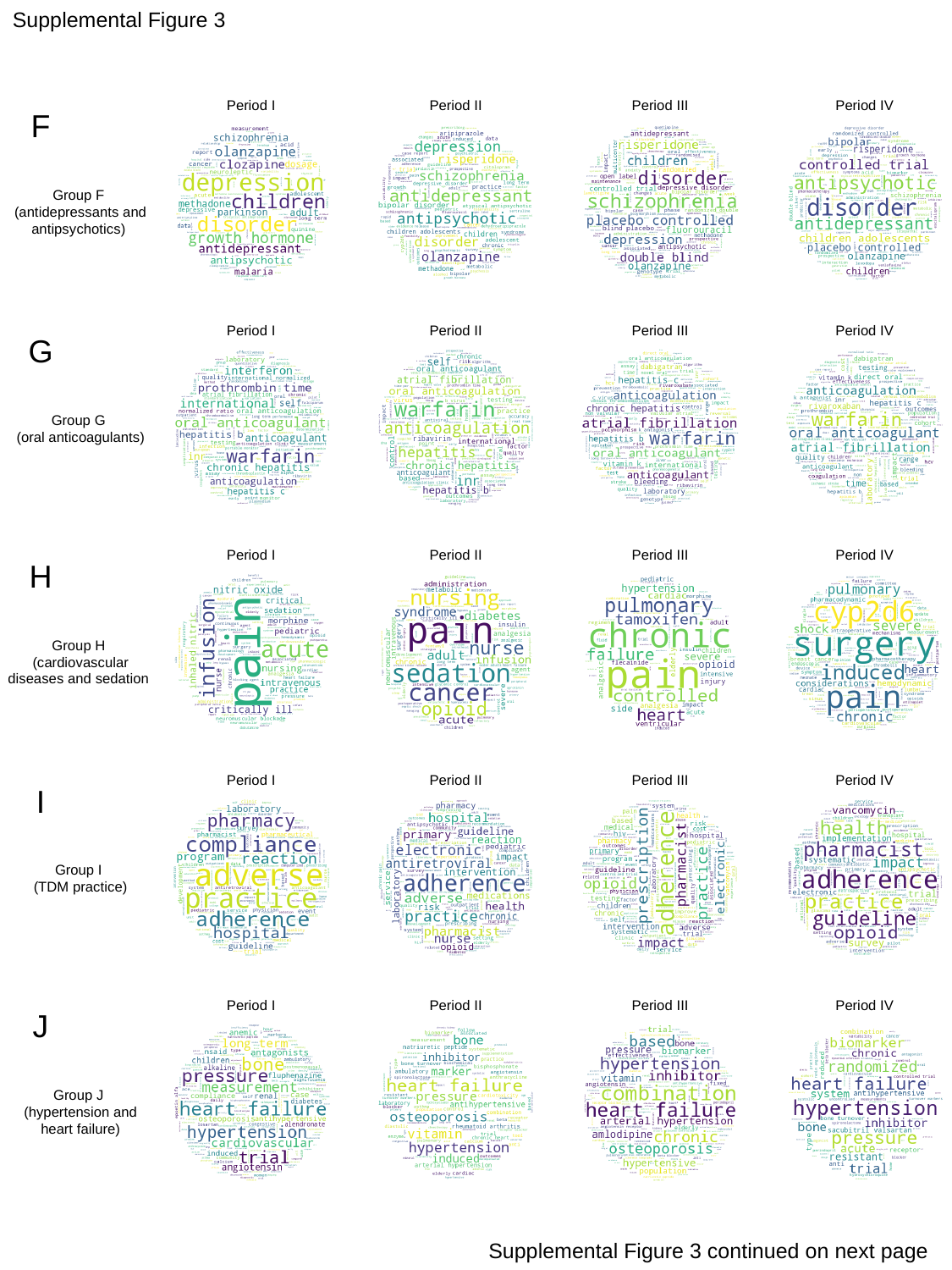

Supplemental Figure 3
Period I
Period II
Period III
Period IV
F
Group F
 (antidepressants and antipsychotics)
Period I
Period II
Period III
Period IV
G
Group G
 (oral anticoagulants)
Period I
Period II
Period III
Period IV
H
Group H
 (cardiovascular diseases and sedation
Period I
Period II
Period III
Period IV
I
Group I
 (TDM practice)
Period I
Period II
Period III
Period IV
J
Group J
 (hypertension and
 heart failure)
Supplemental Figure 3 continued on next page

### Slide 5
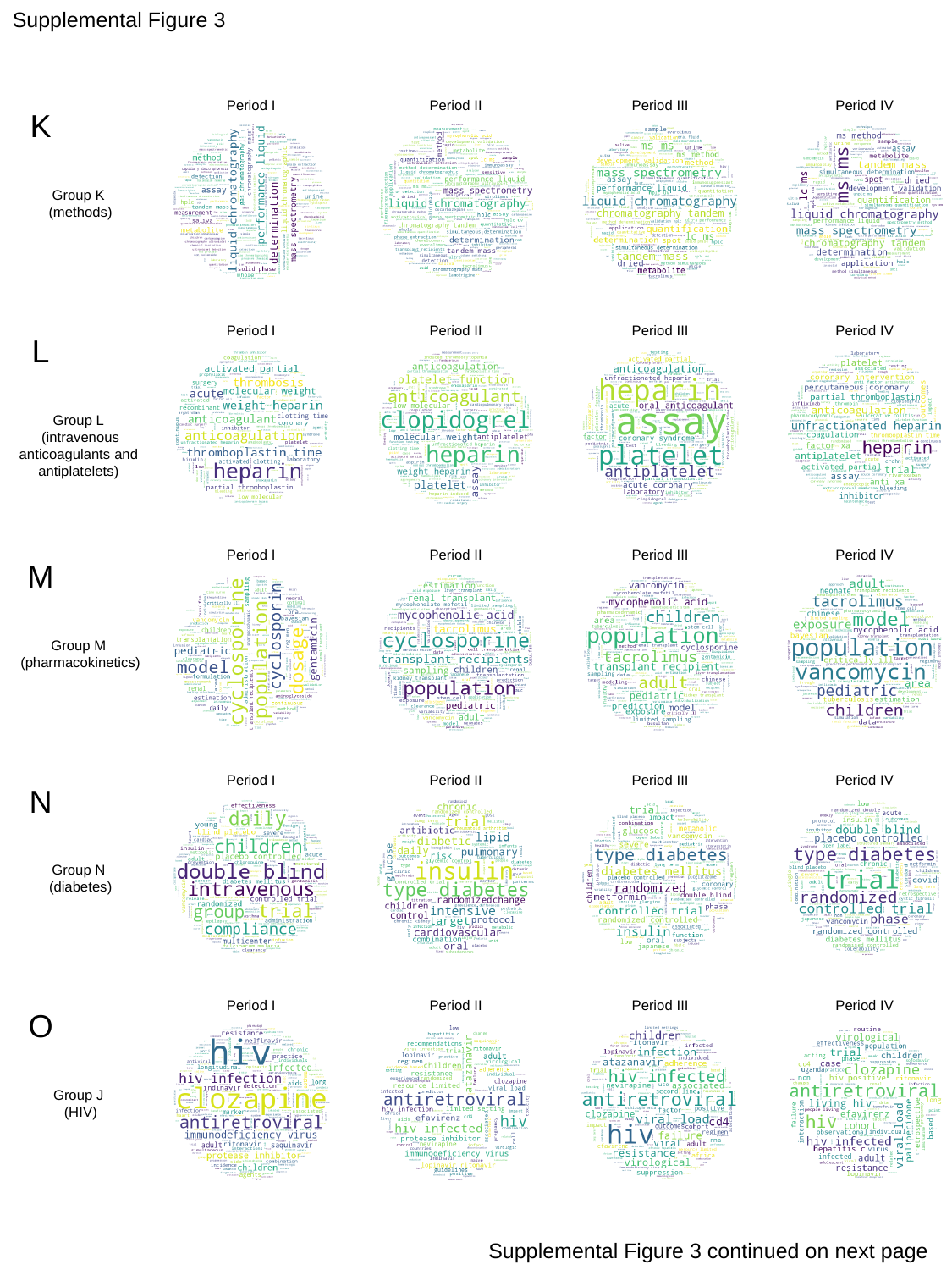

Supplemental Figure 3
Period I
Period II
Period III
Period IV
K
Group K
 (methods)
Period I
Period II
Period III
Period IV
L
Group L
 (intravenous anticoagulants and antiplatelets)
Period I
Period II
Period III
Period IV
M
Group M
 (pharmacokinetics)
Period I
Period II
Period III
Period IV
N
Group N
 (diabetes)
Period I
Period II
Period III
Period IV
O
Group J
 (HIV)
Supplemental Figure 3 continued on next page

### Slide 6
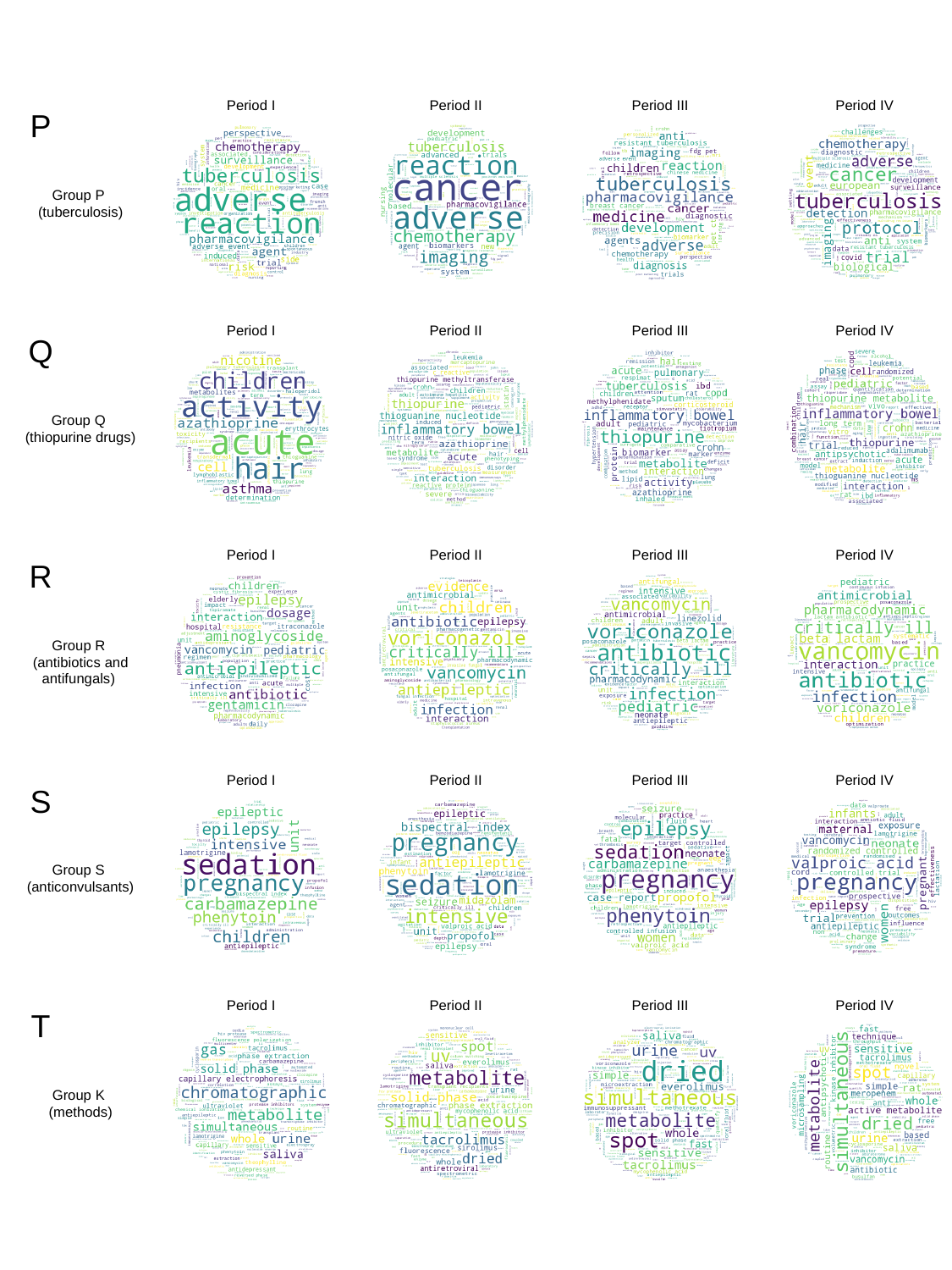

Period I
Period II
Period III
Period IV
P
Group P
 (tuberculosis)
Period I
Period II
Period III
Period IV
Q
Group Q
 (thiopurine drugs)
Period I
Period II
Period III
Period IV
R
Group R
 (antibiotics and antifungals)
Period I
Period II
Period III
Period IV
S
Group S
 (anticonvulsants)
Period I
Period II
Period III
Period IV
T
Group K
 (methods)

### Slide 7
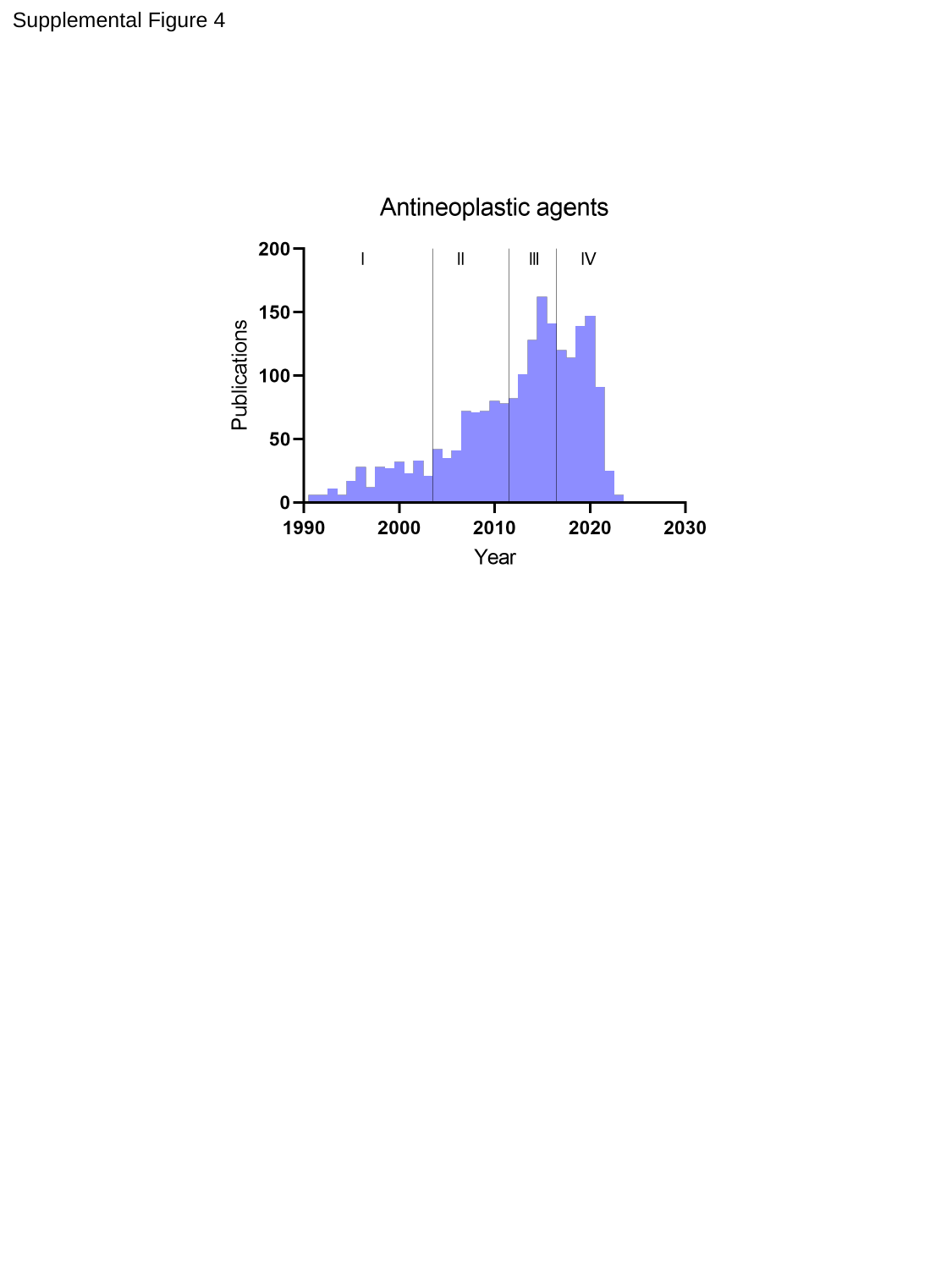

Supplemental Figure 4
